# Associations of white matter hyperintensities with networks of grey matter blood flow and volume in midlife adults: a CARDIA MRI substudy

**DOI:** 10.1101/2021.08.09.21261287

**Authors:** William S.H. Kim, Nicholas J. Luciw, Sarah Atwi, Zahra Shirzadi, Sudipto Dolui, John A. Detre, Ilya M. Nasrallah, Walter Swardfager, R. Nick Bryan, Lenore J. Launer, Bradley J. MacIntosh

**Author notes:** **Corresponding Author:** Bradley J. MacIntosh, Sunnybrook Research Institute, University of Toronto, 2075 Bayview Avenue, Room M6-180, M4N 3M5, Toronto, ON, Canada. **Data Availability** Data in this study are from a sub-sample of men and women who participated in the community-based Coronary Artery Risk Development in Young Adults (CARDIA) brain MRI substudy, examined at the 25-year follow-up exam. Data access is available through the CARDIA Coordinating Center following approval by the CARDIA Publications and Presentations Committee.

## Abstract

White matter hyperintensities (WMHs) are emblematic of cerebral small vessel disease, yet characterization at midlife is poorly studied. Here, we investigated whether WMH volume is associated with brain network alterations in midlife adults. 254 participants from the Coronary Artery Risk Development in Young Adults (CARDIA) study were selected and stratified by WMH burden yielding two groups of equal size (Lo- and Hi-WMH groups). We constructed group-level covariance networks based on cerebral blood flow (CBF) and grey matter volume (GMV) maps across 74 grey matter regions. Through consensus clustering, we found that both CBF and GMV covariance networks were partitioned into modules that were largely consistent between groups. Next, CBF and GMV covariance network topologies were compared between Lo- and Hi-WMH groups at global (clustering coefficient, characteristic path length, global efficiency) and regional (degree, betweenness centrality, local efficiency) levels. At the global level, there were no group differences in either CBF or GMV covariance networks. In contrast, we found group differences in the regional degree, betweenness centrality, and local efficiency of several brain regions in both CBF and GMV covariance networks. Overall, CBF and GMV covariance analyses provide evidence of WMH-related network alterations that were observed at midlife.

## 1. Introduction

White matter hyperintensities (WMHs) of presumed vascular origin are one of the most widely studied markers of cerebral small vessel disease (SVD) (Wardlaw et al., 2019) and are associated with vascular risk factors (de Leeuw et al., 1999), cognitive decline (de Groot et al., 2001), gait abnormalities (Baezner et al., 2008; de Laat et al., 2011), depression (Rabins et al., 1991), and brain atrophy in late-life individuals (Appelman et al., 2009; Godin et al., 2009; Rossi et al., 2006; Schmidt et al., 2005). Whereas WMHs have been well studied in late-life, comparatively less is known regarding their effects on brain function in earlier adult decades of life (Cannistraro et al., 2019). Subclinical WMHs are present in midlife (Bryan et al., 1999; Launer et al., 2015; Wen et al., 2009) and are associated with an increased risk of late-life dementia and early cognitive impairment (d’Arbeloff et al., 2019; Smith et al., 2015).

The mechanisms by which WMHs affect the brain are not fully established, however one potential consequence is the alteration of brain networks (Lawrence et al., 2014). As articulated in a recent review, focal WMHs can impact remote brain regions and structural and functional network connections (ter Telgte et al., 2018). Multivariate methods that integrate information across brain regions may help elucidate the widespread consequences of WMHs beyond conventional neuroimaging biomarker research. SVD has been viewed predominantly as affecting subcortical anatomy proximal to focal lesions, including white matter as well as thalamic and basal ganglia regions (Wardlaw et al., 2013). However, these regions may represent only a proportion of SVD associations across the brain. Covariance analysis of neuroimaging data is one such multivariate approach that can be used to infer network-like associations between brain regions. This technique exploits the phenomenon of inter-region covariation between properties of select brain regions across a population sample (Alexander-Bloch, Giedd, et al., 2013; Melie-García et al., 2013). Typically, covariance analysis constructs group-level networks based upon the pairwise correlation of regional measures of structure such as grey matter volume (GMV) or cortical thickness. These structural covariance networks are thought to arise from developmental coordination or synchronized maturation due to mutually trophic influences (Alexander-Bloch, Raznahan, et al., 2013) and have been shown to partially overlap with both white matter tracts and patterns of functional connectivity (Gong et al., 2012; Kelly et al., 2012). Indeed, WMH volume has been associated with reduced structural covariance network integrity, albeit in older populations (Nestor et al., 2017; Tuladhar et al., 2015).

There is a need for multimodal structural and functional neuroimaging studies to elucidate the mechanisms by which SVD might lead to clinical deficits, especially in earlier adult decades of life (ter Telgte et al., 2018). Cerebral blood flow (CBF) is an important proxy of brain health and metabolism that is amenable to covariance analysis. CBF covariance networks are thought to convey metabolic and vascular information that may be complementary to network analyses performed with structural covariance, blood-oxygenation level-dependent functional MRI (fMRI) or diffusion tensor imaging (DTI) (Luciw et al., 2021; Melie-García et al., 2013).

Graph theory is a principled and data-driven method that can be used to characterize covariance networks and has played a crucial role in establishing the brain as an efficient and sparsely connected “small-world” network (Alexander-Bloch, Giedd, et al., 2013). Considering the brain as a set of nodes (i.e., brain regions) and edges (i.e., connections between nodes, such as pairwise correlations), graph theory produces a number of properties that describe the global and regional topologies of brain networks (Box 1) (Rubinov & Sporns, 2010). Within the context of SVD, the application of graph theory has pointed to anomalous structural (Frey et al., 2020; Lawrence et al., 2014; Reijmer et al., 2016; Tuladhar et al., 2016, 2017) and functional network efficiency (Chen et al., 2019; Sang et al., 2018; Schaefer et al., 2014).

This is Box 1. Global network properties, such as the clustering coefficient (average of the fraction of a node’s neighbors that are neighbors of each other across all nodes), characteristic path length (average shortest path length in the network), and global efficiency (average inverse shortest path length in the network) reflect network-wide attributes. Regional network properties, such as the degree (number of edges on a node), betweenness centrality (measure of the number of shortest paths that travel through a given node), and local efficiency (measure of efficiency in a node’s set of neighbors upon its removal) describe the contribution of individual brain regions that enable efficient communication throughout the network.

This study investigates CBF and GMV covariance networks in midlife adults from the Coronary Artery Risk Development in Young Adults (CARDIA) study. Participants from the 25-year CARDIA brain MRI study were stratified by WMH burden into two groups. CBF and GMV covariance networks were generated for each group and compared using graph theoretical properties of global and regional network topology. We hypothesize that it will be possible to identify differences in CBF and GMV covariance network topology among midlife adults with higher WMH burden when compared against a control group.

## 2. Methods

### 2.1. Participants

The CARDIA study is a longitudinal multi-site prospective study aiming to investigate the evolution of cardiovascular disease over adulthood. Participants were initially recruited in 1985 and between 18 – 30 years of age. The present study uses brain MRI data from the 25-year follow-up exam collected at two of three CARDIA centers in the United States (Minneapolis, MN and Oakland, CA). Participants provided written informed consent at each exam and institutional review boards from each CARDIA center and the coordinating center (University of Minnesota Institutional Review Board, Kaiser-Permanente Northern California Institutional Review Board) approved this study annually.

Clinical measures obtained at the 25-year follow-up exam included: body mass index (BMI, from height and weight); diastolic and systolic blood pressure assessed using a digital blood pressure monitor (OmROn HEM-907XL; Online Fitness, CA); smoking status; diabetes diagnosis (American Diabetes Association, 2011); and blood samples provided estimates of concentrations of high- and low-density lipoprotein cholesterol and triglycerides.

### 2.2. MRI Acquisition

The MRI sequences for the current study were previously described (Launer et al., 2015), and consisted of T1-weighted, T2-weighted fluid-attenuated inversion recovery (FLAIR), and pseudo-continuous arterial spin labeling (ASL) imaging acquired on Siemens 3 Tesla Tim Trio MRI scanners. Isotropic T1-weighted images were acquired in three dimensions using a sagittal MPRAGE sequence (TR/TE/TI = 1900/2.9/900 ms, spatial resolution = 1 mm^3^, FOV = 250 mm, slices = 176, flip angle = 90, GRAPPA = 2, bandwidth = 170 Hz/pixel). Isotropic T2-weighted images were acquired using a sagittal FLAIR sequence (TR/TE/TI = 6000/285/2200 ms, spatial resolution = 1 mm^3^, FOV = 258 mm, slices = 160). CBF maps were calculated from ASL imaging acquired using pseudo-continuous labeling and a two-dimensional multi-slice gradient-echo planar imaging readout (TR/TE = 4000/11 ms, spatial resolution = 3.4 × 3.4 × 5 mm^3^, FOV = 220 mm, flip angle = 90, bandwidth = 3004 Hz/pixel, echo spacing = 0.44 ms, EPI factor = 64, label duration = 1.48 s, offset = 90 mm, radio-frequency pulse gap = 0.36 ms, pulse duration = 0.5 ms, mean z-direction gradient = 0.6 mT/m, post-label delay of 1500 ms [range of 1500 to 2170 ms from most inferior to superior slices], no background suppression, 40 control-label pairs).

### 2.3. MRI Processing

MRI data were processed using SPM8 (www.fil.ion.ucl.ac.uk/spm/software/spm8) and programs developed in MATLAB (MathWorks Inc., Natick, MA). Structural images were processed using a previously described multimodal segmentation algorithm to classify tissue into grey matter (GM), white matter (WM), and cerebrospinal fluid (CSF) (Goldszal et al., 1998; Launer et al., 2015). GM and WM were then assigned to 98 regions using a Talairach-based brain anatomy template (Shen & Davatzikos, 2002). Of these, we chose the 74 regions classified as GM.

WMHs were segmented from structural images using a previously reported deep-learning classification model, built on the U-Net architecture and internal convolutional network Inception ResNet layers (Nasrallah, Pajewski, et al., 2019).

ASL time series data were first motion corrected, followed by regression of residual motion artifacts (Wang, 2012). A Gaussian smoothing kernel with full-width-half-maximum of 5 mm was used to spatially smooth images. A CBF time series was then obtained by pairwise control-label subtraction. CBF image intensities were converted to absolute units after averaging difference images and performing voxel-wise calibration using the ASL control image as the estimate of the equilibrium magnetization (Dolui et al., 2016). CBF maps were then registered to T1-weighted images and mean regional CBF measures were obtained using the same 74 GM regions.

### 2.4. Group Stratification

Participants were selected from an available total sample of 421 for which processed data was fully available. Participants were stratified by WMH burden to create two groups (Figure 1). A nominal WMH volume was measured in all but a small number of participants; as such, we were not powered to stratify groups on the presence alone of WMH volume. Instead, WMH volumes were normalized by intracranial volume and log-transformed to normalize the skewed distribution. Participants in the 7^th^ to 10^th^ deciles of log-transformed WMH volume were denoted as the “Hi-WMH” group (n = 127). For comparison, participants in the 1^st^ to 3^rd^ deciles of log-transformed WMH volume were taken to be the “Lo-WMH” control group (n = 127). The choice to omit the 4^th^ to 6^th^ deciles was to ensure a clear delineation between the two groups.

**Figure 1.**
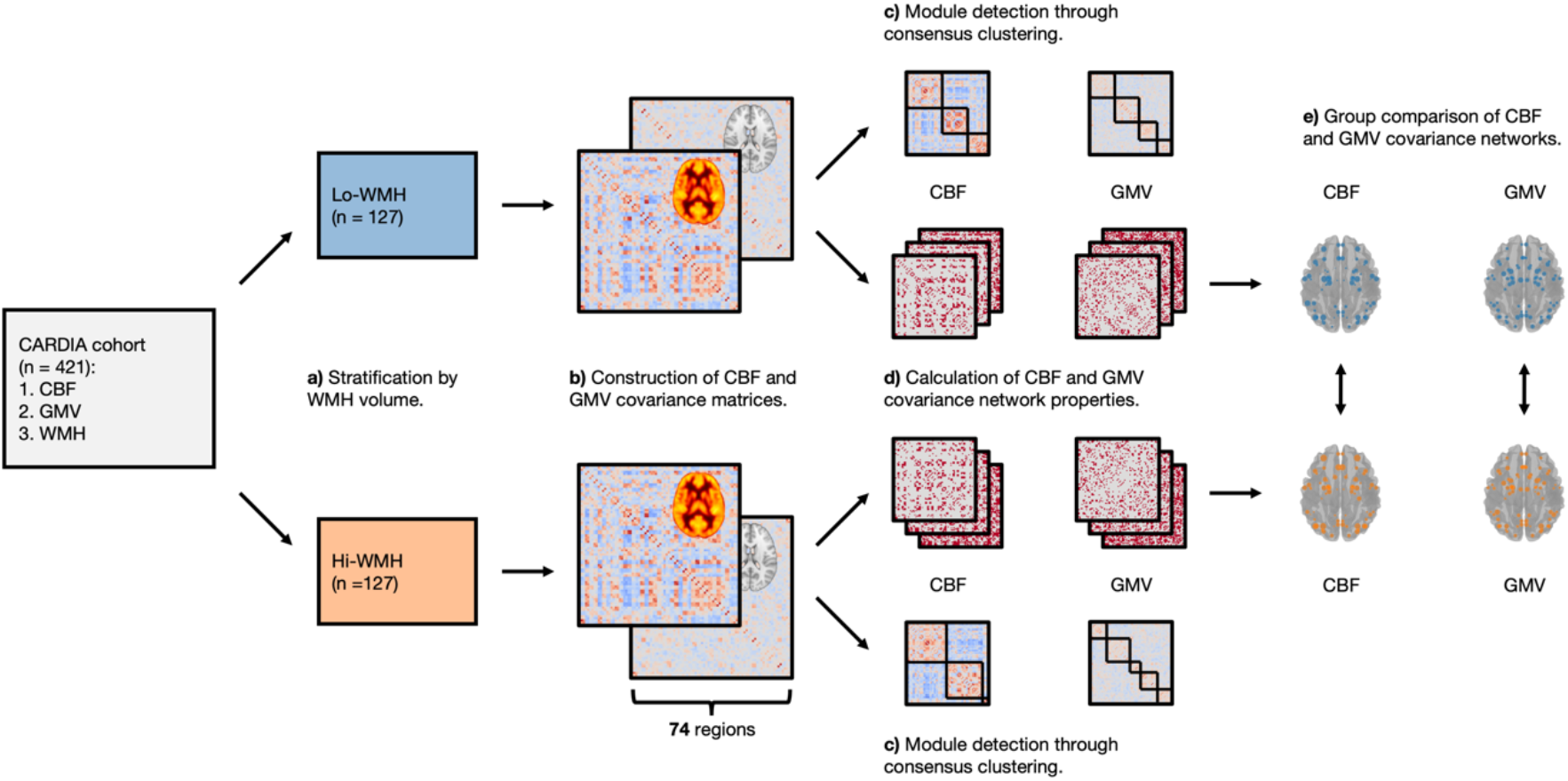
Illustration of analytic workflow. a) Participants were stratified by WMH volume into two groups. b) Group covariance matrices were constructed from CBF and GMV data by Pearson’s correlation coefficient between all brain region pairs, adjusted for age, sex, BMI, race, and MRI site. c) Consensus clustering was performed to detect modules from CBF and GMV covariance matrices. d) Network properties were calculated from CBF and GMV covariance matrices across a range of network densities and area-under-the-curve was integrated to summarize network properties. e) CBF and GMV covariance networks were compared between Lo- and Hi-WMH groups at global (clustering coefficient, characteristic path length, global efficiency) and regional (degree, betweenness centrality, local efficiency) levels using a non-parametric procedure.

### 2.5. Network Analysis

Network analysis was performed in Python 3.7.6 (www.python.org/downloads/release/python-376/) using the Brain Connectivity Toolbox package (Rubinov & Sporns, 2010) (www.pypi.org/project/bctpy/). CBF and GMV measures were intensity normalized and adjusted using a linear regression model for each of the 74 GM brain regions separately by removing the variance attributed to the following variables: age, sex, race, BMI, and MRI site. The adjusted measures were then used to calculate Pearson’s correlation coefficients between all brain region pairs, resulting in 74 × 74 covariance matrices. This procedure was performed for both Lo- and Hi-WMH groups, resulting in a total of four covariance matrices (i.e., two matrices × two groups). Diagonal matrix elements, representing self-connections, were excluded.

We deployed a community detection analysis on the covariance matrices to partition the 74 brain regions into modules (i.e., clusters) with high internal covariance. Briefly, covariance matrices underwent 100 iterations of the Louvain community detection algorithm at selected resolution parameter values of 1.25 for CBF covariance matrices and 0.75 for GMV covariance matrices (see Supplementary Information) (Blondel et al., 2008). Across all iterations, we calculated the probability that a brain region pair was consistently assigned to the same module, yielding a 74 × 74 agreement matrix, which was then thresholded at 0.5 (Cohen & D’Esposito, 2016). Finally, we performed consensus clustering on the agreement matrix using the Louvain algorithm with 100 iterations, yielding a single consensus partition (Lancichinetti & Fortunato, 2012). Unlike conventional clustering approaches, this analytic framework defines the final number of modules from the data.

To characterize network topology, covariance matrices were thresholded and binarized across a range of network densities (0.16 to 0.50, increments of 0.01). The density of a matrix is defined as the number of non-zero edges divided by the total possible number of edges (i.e., [74 × 73] / 2). The minimum network density was chosen such that all brain regions had at least one non-zero edge (i.e., connected to at least one other region within the network). At each of these network densities, we calculated global (clustering coefficient, characteristic path length, and global efficiency) and regional (degree, betweenness centrality, and local efficiency) graph theoretical properties to characterize network topology. Across the range of network densities, global and regional network properties were summarized by the area-under-the-curve (Figure 1).

Finally, we computed the small-world coefficient across the range of network densities, defined as the ratio between normalized clustering coefficient and normalized characteristic path length (i.e., normalized by 100 random networks that preserve the number of nodes and edges as the true network as well as the degree of individual nodes). Networks with a small-world coefficient greater than 1 are thought to maximize efficiency of information transfer while minimizing “wiring costs”, thus providing a balance between functional segregation and integration (Watts & Strogatz, 1998).

### 2.6. Statistical Analysis

Demographic and clinical characteristics were compared between groups using Mann-Whitney U tests for continuous variables and chi-squared tests for categorical variables.

To compare network properties between Lo- and Hi-WMH groups, we used a non-parametric permutation approach with 10,000 permutations. With each permutation, participants were shuffled into two randomized groups of equal size (n = 127, each). CBF and GMV covariance matrices were re-constructed, network properties were re-calculated across the range of network densities, and area-under-the-curve was re-integrated. Differences in network properties between randomized groups were used to create null distributions. The true differences between Lo- and Hi-WMH groups was then compared against the corresponding null distributions, resulting in non-parametric p-values derived as the relative position of the true difference compared to the null distribution. A significance level of 0.05 was chosen for global network properties.

Regional properties were normalized by the corresponding mean of the network before group comparisons. Given the multiple explanatory analyses performed at the regional level, we corrected for multiple comparisons using a false-positive correction, p < (1/N), where N corresponds to the 74 GM regions. We note that this procedure does not strongly control for type I error (Lynall et al., 2010).

Finally, post-hoc analyses investigated regional measures of CBF and GMV between Lo- and Hi-WM groups in regions-of-interest. Independent samples t-tests were used at a significance level of 0.05.

## 3. Results

### 3.1. Demographic & Clinical Characteristics

The Lo- and Hi-WMH groups were matched for most demographic and clinical characteristics, presented in Table 1. There were, however, significant group differences in age (U = 6708.5, p = 0.01), sex (χ^2^ = 12.36, p < 0.001), BMI (U = 7052.5, p = 0.04), and high-density lipoprotein cholesterol (U = 6649.5, p = 0.008). As expected, WMH volume was also different between groups (U = 0.0, p < 0.001).

**Table 1.**
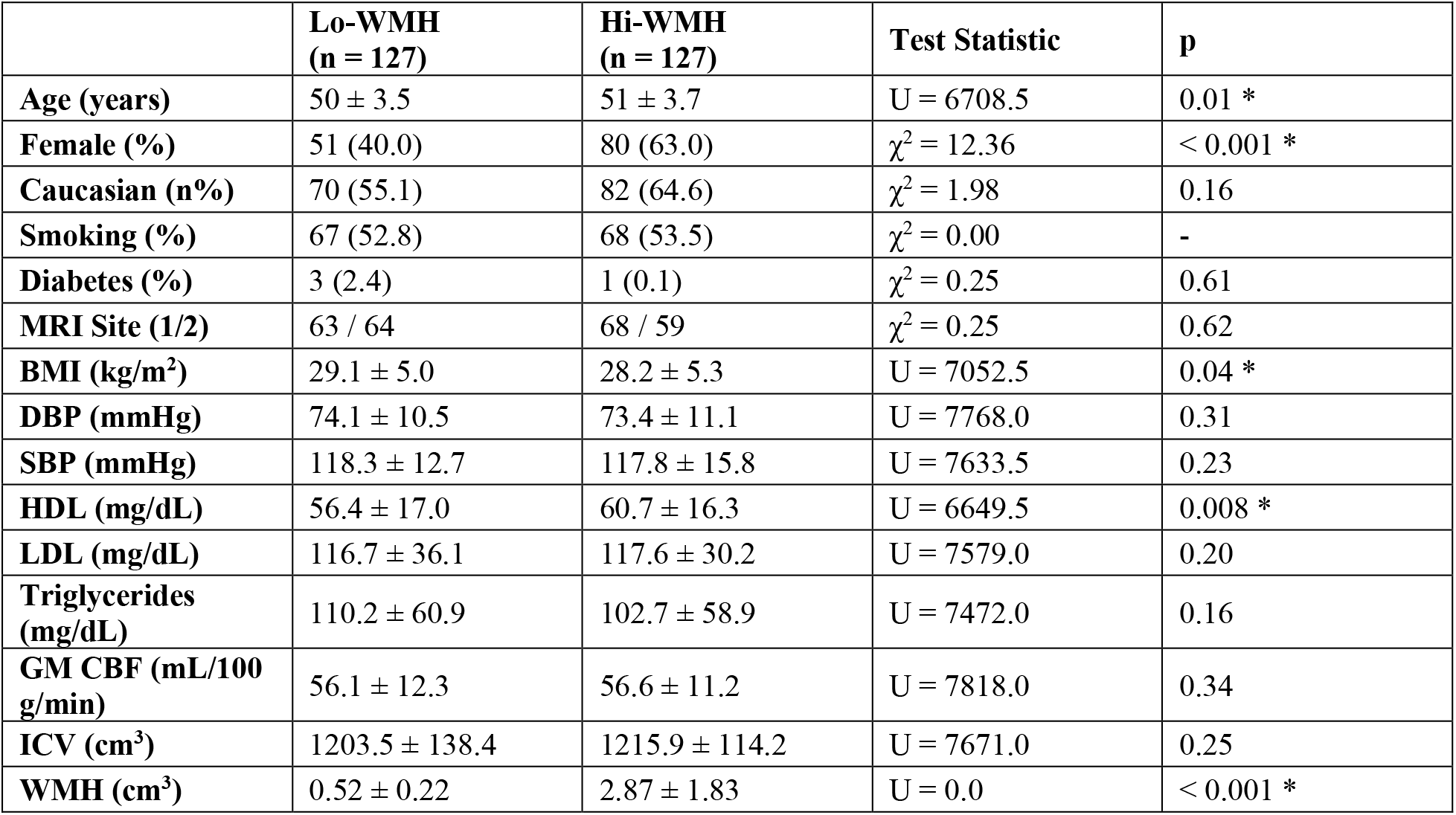
Demographic and clinical characteristics. Data are presented as mean ± standard deviation, or count. p-values were calculated Mann-Whitney U tests for continuous variables and chi-squared tests for categorial variables. * denotes p < 0.05. Abbreviations: BMI, body mass index; DBP, diastolic blood pressure; SBP, systolic blood pressure; HDL, high-density lipoprotein; LDL, low-density lipoprotein; GM, grey matter; ICV, intracranial volume.

### 3.2. Network Modules

Consensus clustering derived three modules from CBF covariance networks that were similar between Lo- and Hi-WMH groups (Figure 2 & Supplementary Table 1). In the Lo-WMH group, Module 1 was the largest and encompassed inferior frontal, temporal, limbic, and subcortical brain regions. Module 2 was comprised of superior and medial frontal, parietal, and occipital brain regions. Module 3 included temporal, occipital, and limbic brain regions as well as the thalami. The Hi-WMH group modules were similar with exception of some discrepancies with the Lo-WMH group that were observed in occipital and temporal regions within Module 3.

**Figure 2.**
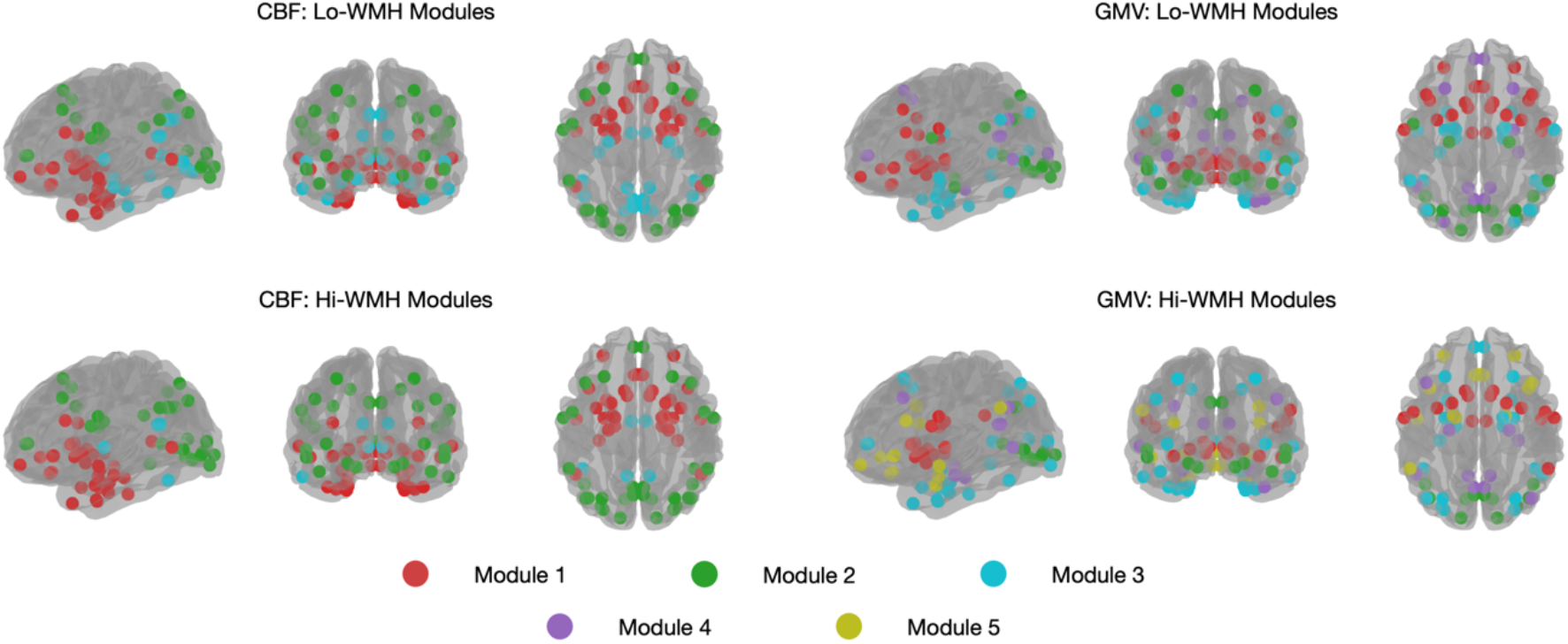
A depiction of the CBF and GMV covariance network modules are shown, as derived from consensus clustering. Parameter resolution values of 1.25 and 0.75 were chosen to generate modules from CBF and GMV covariance matrices using the Louvain community detection algorithm. Consensus clustering derived three modules from CBF covariance networks that were similar between Lo- and Hi-WMH groups. From GMV covariance networks, we observed four and five modules from Lo- and Hi-WMH groups respectively.

From GMV covariance networks, we observed four and five modules from Lo- and Hi-WMH groups, respectively. In the Lo-WMH group, Module 1 consisted of inferior frontal, limbic, parietal, and subcortical brain regions including the thalami. Module 2 consisted of occipital, temporal, and parietal brain regions. Module 3 included limbic, occipital, and temporal brain regions. Module 4 comprised temporal, superior and medial frontal, occipital, and parietal brain regions. In the Hi-WMH group, Module 1 retained subcortical, limbic, and parietal brain regions. Module 3 gained additional frontal and parietal brain regions. Finally, Module 5 consisted of limbic, inferior frontal, parietal, and temporal brain regions.

### 3.3. Small-World Properties

Both CBF and GMV covariance networks from Lo- and Hi-WMH groups exhibited small-world properties across the range of network densities as quantified by normalized clustering coefficients greater than 1 and normalized characteristic path lengths approximately equal to 1 (Figure 3).

**Figure 3.**
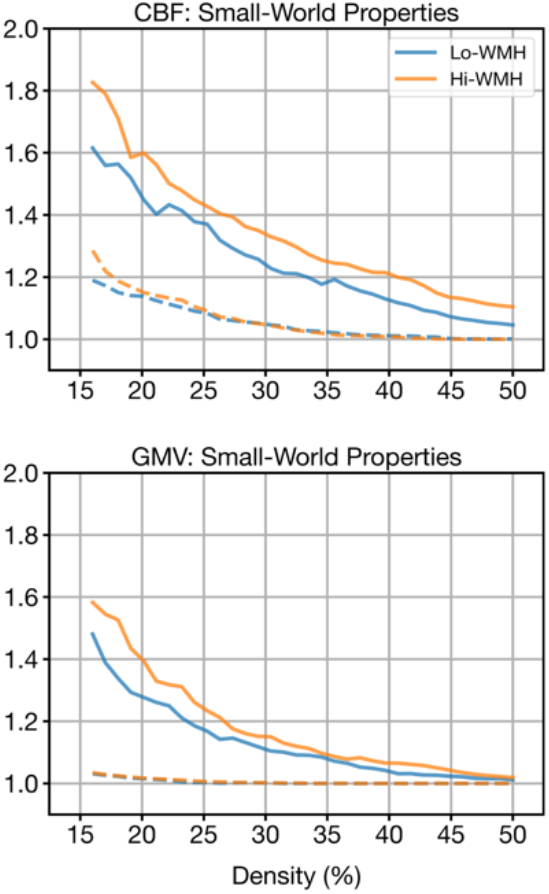
Small-world properties of CBF (top) and GMV (bottom) covariance matrices as a function of network density for Lo- (blue) and Hi-WMH (orange) groups. The small-world coefficient is defined as the ratio between normalized clustering coefficient (solid lines) and normalized characteristic path length (dashed lines). Both Lo- and Hi-WMH groups exhibited small-world topologies (i.e., normalized clustering coefficients greater than 1 and normalized characteristic path lengths approximately equal to 1). True networks were normalized by 100 random networks that preserved the number of nodes and edges as the true network as well as the degree of individual nodes.

### 3.4. Global Network Properties

We found no significant group differences in clustering coefficient, characteristic path length, or global efficiency between CBF covariance networks (clustering coefficient, p = 0.97; characteristic path length, p = 0.92; global efficiency, p = 0.91).

Similarly, there were no group differences between GMV covariance networks (clustering coefficient, p = 0.07; characteristic path length, p = 0.30; global efficiency, p = 0.29) (Figure 4).

**Figure 4.**
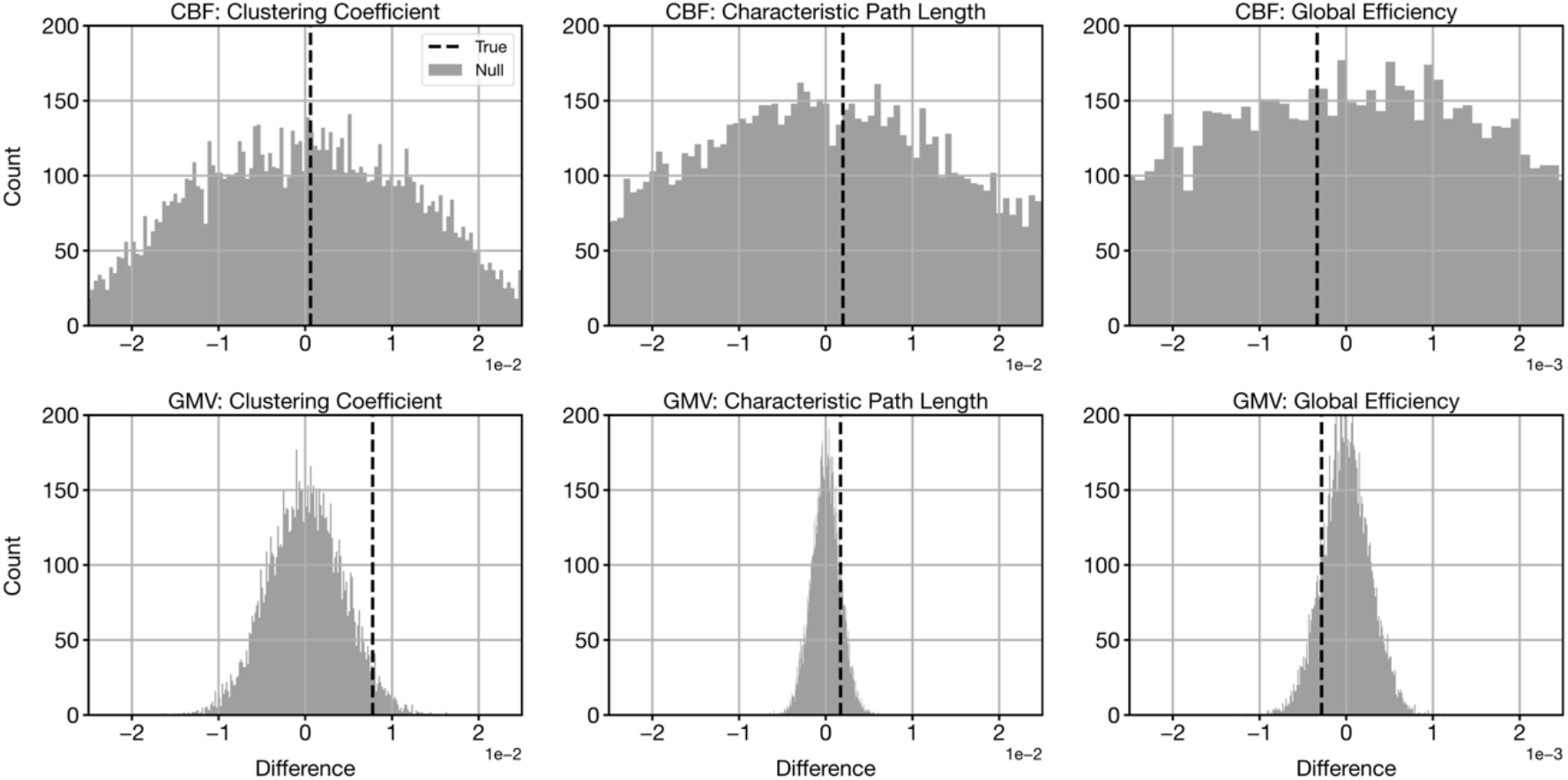
Differences in global network properties in CBF (top row) and GMV (bottom row) covariance networks. Vertical dashed lines indicate true differences between Lo- and Hi-WMH groups (Hi-WMH – Lo-WMH) while histograms illustrate null distributions derived from the non-parametric permutation procedure. There were no significant group differences in any of the global network properties for either CBF or GMV covariance networks at p < 0.05.

### 3.5. Regional Network Properties

Table 2 and Figure 5 present brain regions from CBF and GMV covariance networks with significant differences in degree, betweenness centrality, and local efficiency between Lo- and Hi-WMH groups. Within CBF covariance networks, the Hi-WMH group had lower degree in the right putamen (p = 0.003) relative to the Lo-WMH group. The Hi-WMH group had higher degree (p = 0.002) and betweenness centrality (p = 0.002) in the left nucleus accumbens and higher betweenness centrality in the left cuneus (p = 0.010). We observed no differences in local efficiency within CBF covariance networks.

**Table 2.** Brain regions with significant group differences in regional network properties. The threshold for statistical significance was set at p < 0.0135, after correcting for multiple comparisons. Abbreviations: L, left; R, right.

| Contrast | Region | Network Property | Direction | p |
| --- | --- | --- | --- | --- |
| CBF | R Putamen | Degree | Lo-WMH > Hi-WMH | 0.003 |
|  | L Nucleus Accumbens | Degree | Hi-WMH > Lo-WMH | 0.002 |
|  | L Nucleus Accumbens | Betweenness Centrality | Hi-WMH > Lo-WMH | 0.002 |
|  | L Cuneus | Betweenness Centrality | Hi-WMH > Lo-WMH | 0.010 |
| GMV | L Lingual Gyrus | Degree | Lo-WMH > Hi-WMH | 0.003 |
|  | L Lingual Gyrus | Betweenness Centrality | Lo-WMH > Hi-WMH | 0.004 |
|  | R Lingual Gyrus | Degree | Lo-WMH > Hi-WMH | 0.005 |
|  | R Perirhinal Cortex | Degree | Lo-WMH > Hi-WMH | 0.010 |
|  | L Superior Occipital Gyrus | Local Efficiency | Lo-WMH > Hi-WMH | 0.003 |
|  | L Lateral Occipitotemporal Gyrus | Betweenness Centrality | Hi-WMH > Lo-WMH | 0.010 |
|  | R Superior Parietal Lobule | Local Efficiency | Hi-WMH > Lo-WMH | 0.003 |

**Figure 5.**
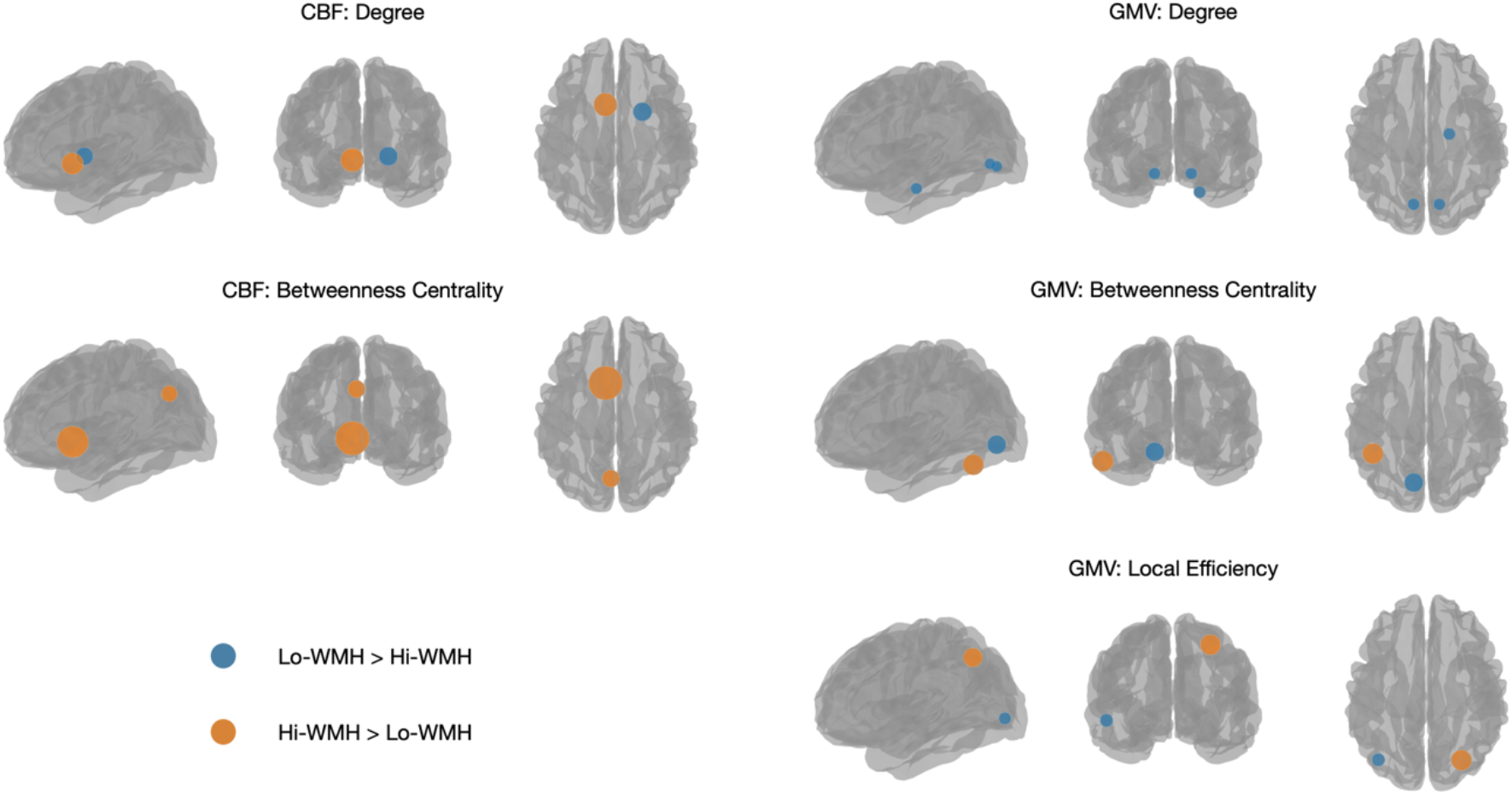
Brain regions that exhibited significant group differences in regional network properties are shown as blue or orange spheres. Blue corresponds to Lo- and Hi-WMH groups, while orange corresponds to Hi-WMH > Lo-WMH. Sphere size corresponds to the magnitude of the group difference. The threshold for statistical significance was set at p < 0.0135.

Within GMV covariance networks, the Hi-WMH group had lower degree (p = 0.003) and betweenness centrality (p = 0.004) in the left lingual gyrus, lower degree (p = 0.005) in the right lingual gyrus, lower degree (p = 0.010) in the right perirhinal cortex, and lower local efficiency (p = 0.003) in the left superior occipital gyrus. The Hi-WMH group had higher betweenness centrality (p = 0.010) in the left lateral occipitotemporal gyrus and higher local efficiency (p = 0.003) in the right superior parietal lobule.

Post-hoc analyses investigating regional measures of CBF and GMV in the above regions of interest revealed no significant group differences (Table 3).

**Table 3.** Post-hoc analyses investigating absolute measures of CBF and GMV in regions-of-interest. Independent samples t-tests were used to compare regional measures of CBF and GMV between Lo- and Hi-WMH groups. No significant differences were observed at a threshold of p < 0.05.

| Contrast | Region | Lo-WMH | Hi-WMH | Test Statistic | p |
| --- | --- | --- | --- | --- | --- |
| CBF | R Putamen | 48.04 ± 10.27 | 47.42 ± 10.34 | t = 0.48 | 0.63 |
|  | L Nucleus Accumbens | 51.08 ± 13.31 | 50.82 ± 14.21 | t = 0.15 | 0.88 |
|  | L Cuneus | 54.62 ± 13.19 | 56.34 ± 13.29 | t = 1.04 | 0.30 |
| GMV | L Lingual Gyrus | 4.05 ± 0.94 | 4.18 ± 0.88 | t = 1.15 | 0.25 |
|  | R Lingual Gyrus | 4.63 ± 1.13 | 4.77 ± 0.92 | t = 1.06 | 0.29 |
|  | R Perirhinal Cortex | 1.56 ± 0.38 | 1.49 ± 0.32 | t = 1.57 | 0.12 |
|  | L Superior Occipital Gyrus | 4.94 ± 1.07 | 4.91 ± 1.03 | t = 0.25 | 0.81 |
|  | L Lateral Occipitotemporal Gyrus | 11.04 ± 1.51 | 11.24 ± 1.43 | t = 1.13 | 0.26 |
|  | R Superior Parietal Lobule | 15.09 ± 2.30 | 14.94 ± 2.21 | t = 0.53 | 0.60 |

## 4. Discussion

In this study, we used graph theory to investigate CBF and GMV covariance networks amongst midlife adults with low vs. high normalized WMH volume. First, we performed a community detection analysis on CBF and GMV covariance networks, resulting in modules of brain regions that were largely consistent between groups. Next, we found that both CBF and GMV covariance networks exhibited small-world topologies across a range of network densities. Finally, we compared CBF and GMV covariance network topologies between groups and found that higher WMH volume was not associated with alterations to global network properties. In contrast, higher WMH volume was associated with altered degree, betweenness centrality, and local efficiency in several brain regions within both CBF and GMV covariance networks. Altogether, these findings provide a holistic picture of physiological and structural patterns across the brain and point to WMH-related network changes in midlife adults.

We first performed community detection analyses to detect modules of brain regions that tend to covary with each other in CBF and GMV covariance networks. CBF covariance matrices were partitioned into three modules that were consistent between Lo- and Hi-WMH groups. Luciw et al. found that communities derived from CBF covariance patterns in adolescents are spatially similar to the brain’s vascular territories (Luciw et al., 2021). Melie-García et al. observed strong CBF covariance between bilateral brain regions in healthy adults (Melie-García et al., 2013). The present study’s findings are similar in both respects, with modules encompassing the brain’s bilateral vascular territories. Meanwhile, GMV covariance matrices were partitioned into four and five modules from Lo- and Hi-WMH groups, respectively. While modules were largely similar between groups, there were notable differences between partitions. For instance, Module 1 (frontal and subcortical) in the Lo-WMH group was divided into Modules 1 (subcortical) and 5 (frontal) in the Hi-WMH group. In the present study, we chose to detect modules for visualization purposes; however, the apparent organization of the brain’s modules suggests alterations to normal covariance patterns as a consequence of higher WMH volume in midlife.

Next, we found that both CBF and GMV covariance networks exhibited small-world topologies, indicating a balance between functional segregation and functional integration (Watts & Strogatz, 1998). Small-world networks are thought to be optimally configured so as to combine clusters of specialized nodes with strategically positioned intermediary edges that minimize overall path lengths, thus improving communication efficiency. These non-random properties have been recapitulated across a wide range of neuroimaging techniques, providing evidence that the brain exhibits a complex yet efficient topology to augment information processing (Bullmore & Sporns, 2009). The current study’s findings further support this hypothesis by demonstrating that networks derived from CBF and GMV covariance analysis similarly obey this organizational structure. Furthermore, higher WMH volume does not disrupt this organization in CBF and GMV covariance networks at midlife.

In comparing Lo- and Hi-WMH groups, we observed no global group differences in the clustering coefficient, characteristic path length, or global efficiency of either CBF or GMV covariance networks. These findings imply that in this sample of midlife adults, the functional integration and segregation of CBF and GMV covariance networks were not associated with higher WMH volume. It is worth noting that in this sample of midlife adults from the CARDIA study, WMH volume is much lower compared to other prospective studies of SVD (Nasrallah, Hsieh, et al., 2019). For instance, other studies reporting on older individuals with comparatively higher WMH volume have found more widespread disruption of structural covariance networks to be associated with higher WMH volume (Nestor et al., 2017). While no global group differences were observed in this cross-sectional study, it may be an accumulation of SVD lesions beginning in midlife that leads to progressive worsening network deficits (Tuladhar et al., 2015).

In contrast, we report significant regional group differences in the degree, betweenness centrality, and local efficiency of several distributed brain regions within both CBF and GMV covariance networks. Namely, we observed CBF covariance alterations in brain regions of the basal ganglia (putamen, nucleus accumbens) and visual (cuneus) networks while GMV covariance revealed alterations in brain regions of the visual (lingual gyri, superior occipital gyrus, lateral occipitotemporal gyrus), limbic (perirhinal cortex), and dorsal attention (superior parietal lobule) networks. The chosen regional network properties are reflective of the importance and influence of individual nodes within a network; in the context of CBF and GMV covariance, these network properties describe the extent to which individual brain regions connect (i.e., covary) with others as well as the impact of these connections on the overall network. The regional and widespread nature of these network alterations, therefore, could reflect subtle WMH-related physiological and structural changes beginning in early stages of disease, and may precede larger network-wide breakdown as reported in more severe stages of SVD (Frey et al., 2020; Lawrence et al., 2014, 2018; Petersen et al., 2020; Reginold et al., 2019; Tuladhar et al., 2016; Xu et al., 2018). Similarly, deficits in regional brain physiology and structure are known to be associated with SVD severity as indexed by increasing WMH volume (Crane et al., 2015; Habes et al., 2016; C. M. Kim et al., 2020; Tuladhar et al., 2015; Tullberg et al., 2004). Multivariate covariance may therefore supplement such univariate analyses in detecting earlier or distinct disease-related alterations (Wee et al., 2013) and in pursuing novel biomarker or hypothesis-generating findings. Notably, our post-hoc analyses examining absolute measures of CBF and GMV in brain regions with significant topological differences revealed no significant group differences.

This study has several limitations. First, by virtue of our study design, we sought to establish group-level covariance networks, which limits our ability to comment on individual participants. While individual-level covariance networks are feasible, it has been shown that individual variability in regional measures may impact subsequent graph theory analyses (H. J. Kim et al., 2016). Second, our group stratification procedure resulted in a decreased sample size. While our analyses retained only a proportion of the original cohort, this procedure resulted in two groups clearly delineated by WMH burden. Third, our false-positive correction does not strongly correct for Type I error. Given the number of regional comparisons made, we chose to avoid an overly conservative correction at the risk of an increased number of false positives. Fourth, the choice of parcellation scheme has been shown to affect brain network estimates (Messé, 2020). Validation studies using external cohort data and alternative parcellation schemes would be desirable to corroborate the findings of the current study. Finally, while our focus was limited to WMHs, it is important to note that other markers of SVD (i.e., enlarged perivascular spaces) were not accounted for and may warrant additional investigation.

In conclusion, we used CBF and GMV covariance analysis to detect WMH-related network changes in midlife adults. We observed modular organization in both CBF and GMV covariance networks that was consistent between Lo- and Hi-WMH groups. We furthermore found that both CBF and GMV covariance networks exhibited small-world properties in both Lo- and Hi-WMH groups, implying an optimal balance between functional integration and segregation. We observed no significant group differences when considering global network properties, suggesting no detectable expression of widespread WMH-related disruptions to functional integration and segregation in this sample of midlife adults. We did, however, detect alterations to regional network properties of CBF and GMV covariance networks in the Hi-WMH group. These findings identify potential WMH-related network alterations in midlife and provide possible avenues for further research of structural and functional brain changes associated with SVD.

## Data Availability

Data in this study are from a sub-sample of men and women who participated in the community-based Coronary Artery Risk Development in Young Adults (CARDIA) brain MRI substudy, examined at the 25-year follow-up exam. Data access is available through the CARDIA Coordinating Center following approval by the CARDIA Publications and Presentations Committee.

## Acknowledgments

The Coronary Artery Risk Development in Young Adults (CARDIA) study is conducted and supported by the National Heart, Lung, and Blood Institute (NHLBI) in collaboration with the University of Alabama at Birmingham (HHSN268201800005I & HHSN268201800007I), Northwestern University (HHSN268201800003I), University of Minnesota (HHSN268201800006I), and Kaiser Foundation Research Institute (HHSN268201800004I). CARDIA was also partially supported by the Intramural Research Program of the National Institute on Aging (NIA) and an intra-agency agreement between NIA and NHLBI (AG0005). This manuscript has been reviewed by CARDIA for scientific content. WSHK received funding from a CIHR CGS-M award. BJM received funding from a NARSAD Independent Investigator Grant from the Brain and Behaviour Research Foundation.

---

**Supplementary Table 1.** CBF and GMV covariance network modules. Consensus clustering derived three modules from CBF covariance networks from Lo- and Hi-WMH groups. From GMV covariance networks, we observed four and five modules from Lo- and Hi-WMH groups, respectively.

| Module | CBF: Lo-WMH | CBF: Hi-WMH | GMV: Lo-WMH | GMV: Hi-WMH |
| --- | --- | --- | --- | --- |
| 1 | L/R Amygdala<br>L/R Caudate<br>L/R Entorhinal Cortex<br>L/R Anterior Cingulate Gyrus<br>L/R Inferior Frontal Gyrus<br>L/R Insula<br>L/R Inferior Temporal Gyrus<br>L/R Lateral Orbitofrontal Gyrus<br>L/R Medial Orbitofrontal Gyrus<br>L/R Middle Temporal Gyrus<br>L/R Nucleus Accumbens<br>L/R Perirhinal Cortex<br>L/R Putamen<br>L/R Superior Temporal Gyrus<br>L/R Temporal Pole<br>L/R Uncus | L/R Amygdala<br>L/R Caudate<br>L/R Entorhinal Cortex<br>L/R Anterior Cingulate Gyrus<br>L/R Hippocampus<br>L/R Inferior Frontal Gyrus<br>L/R Insula<br>L/R Inferior Temporal Gyrus<br>L/R Lateral Orbitofrontal Gyrus<br>L/R Medial Orbitofrontal Gyrus<br>L/R Middle Temporal Gyrus<br>L/R Nucleus Accumbens<br>L/R Perirhinal Cortex<br>L/R Parahippocampal Gyrus<br>L/R Putamen<br>L/R Superior Temporal Gyrus<br>L/R Temporal Pole<br>L/R Uncus | L/R Caudate<br>L/R Anterior Cingulate Gyrus<br>L/R Inferior Frontal Gyrus<br>L/R Insula<br>L/R Lateral Orbitofrontal Gyrus<br>L/R Medial Orbitofrontal Gyrus<br>L/R Middle Frontal Gyrus<br>L/R Nucleus Accumbens<br>L Postcentral Gyrus<br>R Precentral Gyrus<br>L/R Putamen<br>R Superior Temporal Gyrus<br>L/R Thalamus | L/R Caudate<br>L/R Insula<br>L/R Nucleus Accumbens<br>L/R Postcentral Gyrus<br>L/R Precentral Gyrus<br>L/R Putamen<br>R Superior Temporal Gyrus<br>L/R Thalamus |
| 2 | L/R Angular Gyrus<br>L/R Inferior Occipital Gyrus<br>L/R Medial Frontal Gyrus<br>L/R Middle Frontal Gyrus<br>L/R Middle Occipital Gyrus<br>L/R Occipital Pole<br>L/R Postcentral Gyrus<br>L/R Precentral Gyrus<br>L/R Superior Frontal Gyrus<br>L/R Supramarginal Gyrus<br>L/R Superior Occipital Gyrus<br>L/R Superior Parietal Lobule | L/R Angular Gyrus<br>L/R Cuneus<br>L/R Inferior Occipital Gyrus<br>L/R Lingual Gyrus<br>L/R Lateral Occipitotemporal Gyrus<br>L/R Medial Frontal Gyrus<br>L/R Middle Frontal Gyrus<br>L/R Middle Occipital Gyrus<br>L/R Medial Occipitotemporal Gyrus<br>L/R Occipital Pole<br>L/R Postcentral Gyrus<br>L/R Precuneus<br>L/R Precentral Gyrus<br>L/R Superior Frontal Gyrus<br>L/R Supramarginal Gyrus<br>L/R Superior Occipital Gyrus<br>L/R Superior Parietal Lobule | L/R Cuneus<br>L/R Hippocampus<br>L/R Inferior Occipital Gyrus<br>L/R Lingual Gyrus<br>L/R Medial Occipitotemporal Gyrus<br>L/R Occipital Pole<br>L/R Precentral Gyrus<br>R Supramarginal Gyrus<br>L Superior Occipital Gyrus<br>L/R Superior Parietal Lobule | L/R Cuneus<br>R Inferior Occipital Gyrus<br>L/R Lingual Gyrus<br>L/R Medial Occipitotemporal Gyrus<br>L/R Occipital Pole<br>L/R Superior Occipital Gyrus |
| 3 | L/R Cuneus<br>L/R Hippocampus<br>L/R Lingual Gyrus<br>L/R Lateral Occipitotemporal Gyrus<br>L/R Medial Occipitotemporal Gyrus<br>L/R Posterior Cingulate Gyrus<br>L/R Parahippocampal Gyrus<br>L/R Precuneus<br>L/R Thalamus | L/R Posterior Cingulate Gyrus<br>L/R Thalamus | L/R Amygdala<br>L/R Angular Gyrus<br>L/R Entorhinal Cortex<br>L Inferior Temporal Gyrus<br>L/R Lateral Occipitotemporal Gyrus<br>R Middle Occipital Gyrus<br>R Middle Temporal Gyrus<br>R Postcentral Gyrus<br>L/R Perirhinal Cortex<br>L Parahippocampal Gyrus<br>L Supramarginal Gyrus<br>R Superior Occipital Gyrus<br>L Superior Temporal Gyrus<br>L/R Temporal Pole<br>L/R Uncus | R Amygdala<br>L Angular Gyrus<br>L/R Entorhinal Cortex<br>L Inferior Occipital Gyrus<br>L/R Inferior Temporal Gyrus<br>L/R Lateral Occipitotemporal Gyrus<br>L/R Medial Frontal Gyrus<br>L/R Middle Occipital Gyrus<br>R Middle Temporal Gyrus<br>L/R Perirhinal Cortex<br>L Parahippocampal Gyrus<br>L/R Superior Frontal Gyrus<br>L/R Superior Parietal Lobule<br>L/R Temporal Pole |
| 4 |  |  | R Inferior Temporal Gyrus<br>L/R Medial Frontal Gyrus<br>L Occipital Gyrus<br>L Middle Temporal Gyrus<br>L/R Posterior Cingulate Gyrus<br>R Parahippocampal Gyrus<br>L/R Precuneus<br>L/R Superior Frontal Gyrus | R Angular Gyrus<br>L/R Hippocampus<br>L Middle Frontal Gyrus<br>L Middle Temporal Gyrus<br>L/R Posterior Cingulate Gyrus<br>R Parahippocampal Gyrus<br>L/R Precuneus |
| 5 |  |  |  | L Amygdala<br>L/R Anterior Cingulate Gyrus<br>L/R Inferior Frontal Gyrus<br>L/R Lateral Orbitofrontal Gyrus<br>L/R Medial Orbitofrontal Gyrus<br>R Middle Frontal Gyrus<br>L Supramarginal Gyrus<br>L Superior Temporal Gyrus<br>L/R Uncus |

## Supplementary Information

One methodological issue of community detection analysis is the so-called resolution limit, which describes an algorithm’s inability to identify modules below a certain size (Sporns & Betzel, 2016). To address this, we assessed the stability of the Louvain algorithm by varying the resolution parameter, a variable that dictates the size and number of partitioned modules. Across a range of resolution parameter values (0.5 to 1.5, increments of 0.05), we performed 1,000 iterations of the Louvain algorithm. The stability of the resulting partitions at each resolution parameter value was assessed by calculating the mean Hamming distance across all partition pairs. The resolution parameter value corresponding to the lowest (i.e., most stable) mean Hamming distance was then used for subsequent consensus clustering. This procedure was performed separately for GMV and CBF covariance matrices, constructed across all participants.

## References

Alexander-Bloch, A., Giedd, J. N., & Bullmore, E. (2013). Imaging structural co-variance between human brain regions. In Nature Reviews Neuroscience. https://doi.org/10.1038/nrn3465

Alexander-Bloch, A., Raznahan, A., Bullmore, E., & Giedd, J. (2013). The convergence of maturational change and structural covariance in human cortical networks. Journal of Neuroscience. https://doi.org/10.1523/JNEUROSCI.3554-12.2013

American Diabetes Association. (2011). Diagnosis and classification of diabetes mellitus. In Diabetes Care. https://doi.org/10.2337/dc11-S062

Appelman, A. P. A., Exalto, L. G., van der Graaf, Y., Biessels, G. J., Mali, W. P. T. M., & Geerlings, M. I. (2009). White matter lesions and brain atrophy: More than shared risk factors? A systematic review. In Cerebrovascular Diseases. https://doi.org/10.1159/000226774

Baezner, H., Blahak, C., Poggesi, A., Pantoni, L., Inzitari, D., Chabriat, H., Erkinjuntti, T., Fazekas, F., Ferro, J. M., Langhorne, P., O’Brien, J., Scheltens, P., Visser, M. C., Wahlund, L. O., Waldemar, G., Wallin, A., & Hennerici, M. G. (2008). Association of gait and balance disorders with age-related white matter changes: The LADIS Study. Neurology. https://doi.org/10.1212/01.wnl.0000305959.46197.e6

Blondel, V. D., Guillaume, J. L., Lambiotte, R., & Lefebvre, E. (2008). Fast unfolding of communities in large networks. Journal of Statistical Mechanics: Theory and Experiment, 2008(10), 10008. https://doi.org/10.1088/1742-5468/2008/10/P10008

Bryan, R. N., Cai, J., Burke, G., Hutchinson, R. G., Liao, D., Toole, J. F., Dagher, A. P., & Cooper, L. (1999). Prevalence and anatomic characteristics of infarct-like lesions on MR images of middle-aged adults: The atherosclerosis risk in communities study. American Journal of Neuroradiology.

Bullmore, E., & Sporns, O. (2009). Complex brain networks: Graph theoretical analysis of structural and functional systems. In Nature Reviews Neuroscience. https://doi.org/10.1038/nrn2575

Cannistraro, R. J., Badi, M., Eidelman, B. H., Dickson, D. W., Middlebrooks, E. H., & Meschia, J. F. (2019). CNS small vessel disease: A clinical review. In Neurology. https://doi.org/10.1212/WNL.0000000000007654

Chen, H., Huang, L., Yang, D., Ye, Q., Guo, M., Qin, R., Luo, C., Li, M., Ye, L., Zhang, B., & Xu, Y. (2019). Nodal Global Efficiency in Front-Parietal Lobe Mediated Periventricular White Matter Hyperintensity (PWMH)-Related Cognitive Impairment. Frontiers in Aging Neuroscience. https://doi.org/10.3389/fnagi.2019.00347

Cohen, J. R., & D’Esposito, M. (2016). The segregation and integration of distinct brain networks and their relationship to cognition. Journal of Neuroscience, 36(48), 12083– 12094. https://doi.org/10.1523/JNEUROSCI.2965-15.2016

Crane, D. E., Black, S. E., Ganda, A., Mikulis, D. J., Nestor, S. M., Donahue, M. J., & MacIntosh, B. J. (2015). Grey matter blood flow and volume are reduced in association with white matter hyperintensity lesion burden: A cross-sectional MRI study. Frontiers in Aging Neuroscience. https://doi.org/10.3389/fnagi.2015.00131

d’Arbeloff, T., Elliott, M. L., Knodt, A. R., Melzer, T. R., Keenan, R., Ireland, D., Ramrakha, S., Poulton, R., Anderson, T., Caspi, A., Moffitt, T. E., & Hariri, A. R. (2019). White matter hyperintensities are common in midlife and already associated with cognitive decline. Brain Communications. https://doi.org/10.1093/braincomms/fcz041

de Groot, J. C., de Leeuw, F. E., Oudkerk, M., Hofman, A., Jolles, J., & Breteler, M. M. B. (2001). Cerebral white matter lesions and subjective cognitive dysfunction: The Rotterdam scan study. Neurology. https://doi.org/10.1212/WNL.56.11.1539

de Laat, K. F., Tuladhar, A. M., van Norden, A. G. W., Norris, D. G., Zwiers, M. P., & de Leeuw, F. E. (2011). Loss of white matter integrity is associated with gait disorders in cerebral small vessel disease. Brain. https://doi.org/10.1093/brain/awq343

de Leeuw, F. E., de Groot, J. C., Oudkerk, M., Witteman, J. C. M., Hofman, A., van Gijn, J., & Breteler, M. M. B. (1999). A follow-up study of blood pressure and cerebral white matter lesions. Annals of Neurology. https://doi.org/10.1002/1531-8249(199912)46:6<827::AID-ANA4>3.0.CO;2-H

Dolui, S., Wang, Z., Wang, D. J. J., Mattay, R., Finkel, M., Elliott, M., Desiderio, L., Inglis, B., Mueller, B., Stafford, R. B., Launer, L. J., Jacobs, D. R., Bryan, R. N., & Detre, J. A. (2016). Comparison of non-invasive MRI measurements of cerebral blood flow in a large multisite cohort. Journal of Cerebral Blood Flow and Metabolism. https://doi.org/10.1177/0271678X16646124

Frey, B. M., Petersen, M., Schlemm, E., Mayer, C., Hanning, U., Engelke, K., Fiehler, J., Borof, K., Jagodzinski, A., Gerloff, C., Thomalla, G., & Cheng, B. (2020). White matter integrity and structural brain network topology in cerebral small vessel disease: The Hamburg city health study. Human Brain Mapping, 42(5), 1406–1415. https://doi.org/10.1002/hbm.25301

Godin, O., Maillard, P., Crivello, F., Alpérovitch, A., Mazoyer, B., Tzourio, C., & Dufouil, C. (2009). Association of white-matter lesions with brain atrophy markers: The three-city Dijon MRI study. Cerebrovascular Diseases. https://doi.org/10.1159/000226117

Goldszal, A. F., Davatzikos, C., Pham, D. L., Yan, M. X. H., Bryan, R. N., & Resnick, S. M. (1998). An image-processing system for qualitative and quantitative volumetric analysis of brain images. Journal of Computer Assisted Tomography. https://doi.org/10.1097/00004728-199809000-00030

Gong, G., He, Y., Chen, Z. J., & Evans, A. C. (2012). Convergence and divergence of thickness correlations with diffusion connections across the human cerebral cortex. NeuroImage. https://doi.org/10.1016/j.neuroimage.2011.08.017

Habes, M., Erus, G., Toledo, J. B., Zhang, T., Bryan, N., Launer, L. J., Rosseel, Y., Janowitz, D., Doshi, J., Van Der Auwera, S., Von Sarnowski, B., Hegenscheid, K., Hosten, N., Homuth, G., Völzke, H., Schminke, U., Hoffmann, W., Grabe, H. J., & Davatzikos, C. (2016). White matter hyperintensities and imaging patterns of brain ageing in the general population. Brain, 139(4), 1164–1179. https://doi.org/10.1093/brain/aww008

Kelly, C., Toro, R., di Martino, A., Cox, C. L., Bellec, P., Castellanos, F. X., & Milham, M. P. (2012). A convergent functional architecture of the insula emerges across imaging modalities. NeuroImage. https://doi.org/10.1016/j.neuroimage.2012.03.021

Kim, C. M., Alvarado, R. L., Stephens, K., Wey, H. Y., Wang, D. J. J., Leritz, E. C., & Salat, D. H. (2020). Associations between cerebral blood flow and structural and functional brain imaging measures in individuals with neuropsychologically defined mild cognitive impairment. Neurobiology of Aging, 86, 64–74. https://doi.org/10.1016/j.neurobiolaging.2019.10.023

Kim, H. J., Shin, J. H., Han, C. E., Kim, H. J., Na, D. L., Seo, S. W., & Seong, J. K. (2016). Using individualized brain network for analyzing structural covariance of the cerebral cortex in Alzheimer’s patients. Frontiers in Neuroscience. https://doi.org/10.3389/fnins.2016.00394

Lancichinetti, A., & Fortunato, S. (2012). Consensus clustering in complex networks. Scientific Reports, 2(1), 1–7. https://doi.org/10.1038/srep00336

Launer, L. J., Lewis, C. E., Schreiner, P. J., Sidney, S., Battapady, H., Jacobs, D. R., Lim, K. O., D’Esposito, M., Zhang, Q., Reis, J., Davatzikos, C., & Bryan, R. N. (2015). Vascular factors and multiple measures of early brain health: CARDIA brain MRI study. PLoS ONE. https://doi.org/10.1371/journal.pone.0122138

Lawrence, A. J., Chung, A. W., Morris, R. G., Markus, H. S., & Barrick, T. R. (2014). Structural network efficiency is associated with cognitive impairment in small-vessel disease. Neurology. https://doi.org/10.1212/WNL.0000000000000612

Lawrence, A. J., Zeestraten, E. A., Benjamin, P., Lambert, C. P., Morris, R. G., Barrick, T. R., & Markus, H. S. (2018). Longitudinal decline in structural networks predicts dementia in cerebral small vessel disease. Neurology, 90(21), e1898–e1910. https://doi.org/10.1212/WNL.0000000000005551

Luciw, N. J., Toma, S., Goldstein, B. I., & MacIntosh, B. J. (2021). Correspondence between patterns of cerebral blood flow and structure in adolescents with and without bipolar disorder. Journal of Cerebral Blood Flow & Metabolism. https://doi.org/10.1177/0271678X21989246.

Lynall, M. E., Bassett, D. S., Kerwin, R., McKenna, P. J., Kitzbichler, M., Muller, U., & Bullmore, E. (2010). Functional connectivity and brain networks in schizophrenia. Journal of Neuroscience. https://doi.org/10.1523/JNEUROSCI.0333-10.2010

Melie-García, L., Sanabria-Diaz, G., & Sánchez-Catasús, C. (2013). Studying the topological organization of the cerebral blood flow fluctuations in resting state. NeuroImage. https://doi.org/10.1016/j.neuroimage.2012.08.082

Messé, A. (2020). Parcellation influence on the connectivity-based structure–function relationship in the human brain. Human Brain Mapping. https://doi.org/10.1002/hbm.24866

Nasrallah, I. M., Hsieh, M. K., Erus, G., Battapady, H., Dolui, S., Detre, J. A., Launer, L. J., Jacobs, D. R., Davatzikos, C., & Bryan, R. N. (2019). White matter lesion penumbra shows abnormalities on structural and physiologic MRIs in the coronary artery risk development in young adults cohort. American Journal of Neuroradiology. https://doi.org/10.3174/ajnr.A6119

Nasrallah, I. M., Pajewski, N. M., Auchus, Al. P., Chelune, G., Cheung, A. K., Cleveland, M. L., Coker, L. H., Crowe, M. G., Cushman, W. C., Cutler, J. A., Davatzikos, C., Desiderio, L., Doshi, J., Erus, G., Fine, L. J., Gaussoin, S. A., Harris, D., Johnson, K. C., Kimmel, P. L., … Bryan, R. N. (2019). Association of intensive vs standard blood pressure control with cerebral white matter lesions. JAMA - Journal of the American Medical Association. https://doi.org/10.1001/jama.2019.10551

Nestor, S. M., Mišić, B., Ramirez, J., Zhao, J., Graham, S. J., Verhoeff, N. P. L. G., Stuss, D. T., Masellis, M., & Black, S. E. (2017). Small vessel disease is linked to disrupted structural network covariance in Alzheimer’s disease. Alzheimer’s and Dementia. https://doi.org/10.1016/j.jalz.2016.12.007

Petersen, M., Frey, B. M., Schlemm, E., Mayer, C., Hanning, U., Engelke, K., Fiehler, J., Borof, K., Jagodzinski, A., Gerloff, C., Thomalla, G., & Cheng, B. (2020). Network Localisation of White Matter Damage in Cerebral Small Vessel Disease. Scientific Reports, 10(1). https://doi.org/10.1038/s41598-020-66013-w

Rabins, P. v., Pearlson, G. D., Aylward, E., Kumar, A. J., & Dowell, K. (1991). Cortical magnetic resonance imaging changes in elderly inpatients with major depression. American Journal of Psychiatry. https://doi.org/10.1176/ajp.148.5.617

Reginold, W., Sam, K., Poublanc, J., Fisher, J., Crawley, A., & Mikulis, D. J. (2019). The efficiency of the brain connectome is associated with cerebrovascular reactivity in persons with white matter hyperintensities. Human Brain Mapping, 40(12), 3647–3656. https://doi.org/10.1002/hbm.24622

Reijmer, Y. D., Fotiadis, P., Piantoni, G., Boulouis, G., Kelly, K. E., Gurol, M. E., Leemans, A., O’Sullivan, M. J., Greenberg, S. M., & Viswanathan, A. (2016). Small vessel disease and cognitive impairment: The relevance of central network connections. Human Brain Mapping. https://doi.org/10.1002/hbm.23186

Rossi, R., Boccardi, M., Sabattoli, F., Galluzzi, S., Alaimo, G., Testa, C., & Frisoni, G. B. (2006). Topographic correspondence between white matter hyperintensities and brain atrophy. Journal of Neurology. https://doi.org/10.1007/s00415-006-0133-z

Rubinov, M., & Sporns, O. (2010). Complex network measures of brain connectivity: Uses and interpretations. NeuroImage. https://doi.org/10.1016/j.neuroimage.2009.10.003

Sang, L., Chen, L., Wang, L., Zhang, J., Zhang, Y., Li, P., Li, C., & Qiu, M. (2018). Progressively disrupted brain functional connectivity network in subcortical ischemic vascular cognitive impairment patients. Frontiers in Neurology. https://doi.org/10.3389/fneur.2018.00094

Schaefer, A., Quinque, E. M., Kipping, J. A., Arélin, K., Roggenhofer, E., Frisch, S., Villringer, A., Mueller, K., & Schroeter, M. L. (2014). Early small vessel disease affects frontoparietal and cerebellar hubs in close correlation with clinical symptoms -a resting-state fMRI study. Journal of Cerebral Blood Flow and Metabolism. https://doi.org/10.1038/jcbfm.2014.70

Schmidt, R., Ropele, S., Enzinger, C., Petrovic, K., Smith, S., Schmidt, H., Matthews, P. M., & Fazekas, F. (2005). White matter lesion progression, brain atrophy, and cognitive decline: The Austrian stroke prevention study. Annals of Neurology. https://doi.org/10.1002/ana.20630

Shen, D., & Davatzikos, C. (2002). HAMMER: Hierarchical attribute matching mechanism for elastic registration. IEEE Transactions on Medical Imaging, 21(11), 1421–1439. https://doi.org/10.1109/TMI.2002.803111

Smith, E. E., O’Donnell, M., Dagenais, G., Lear, S. A., Wielgosz, A., Sharma, M., Poirier, P., Stotts, G., Black, S. E., Strother, S., Noseworthy, M. D., Benavente, O., Modi, J., Goyal, M., Batool, S., Sanchez, K., Hill, V., McCreary, C. R., Frayne, R., … Yusuf, S. (2015). Early cerebral small vessel disease and brain volume, cognition, and gait. Annals of Neurology. https://doi.org/10.1002/ana.24320

Sporns, O., & Betzel, R. F. (2016). Modular brain networks. Annual Review of Psychology, 67, 613–640. https://doi.org/10.1146/annurev-psych-122414-033634

ter Telgte, A., van Leijsen, E. M. C., Wiegertjes, K., Klijn, C. J. M., Tuladhar, A. M., & de Leeuw, F. E. (2018). Cerebral small vessel disease: From a focal to a global perspective. In Nature Reviews Neurology. https://doi.org/10.1038/s41582-018-0014-y

Tuladhar, A. M., Lawrence, A., Norris, D. G., Barrick, T. R., Markus, H. S., & de Leeuw, F. E. (2017). Disruption of rich club organisation in cerebral small vessel disease. Human Brain Mapping. https://doi.org/10.1002/hbm.23479

Tuladhar, A. M., Reid, A. T., Shumskaya, E., de Laat, K. F., van Norden, A. G. W., van Dijk, E. J., Norris, D. G., & de Leeuw, F. E. (2015). Relationship between white matter hyperintensities, cortical thickness, and cognition. Stroke. https://doi.org/10.1161/STROKEAHA.114.007146

Tuladhar, A. M., van Dijk, E., Zwiers, M. P., van Norden, A. G. W., de Laat, K. F., Shumskaya, E., Norris, D. G., & de Leeuw, F. E. (2016). Structural network connectivity and cognition in cerebral small vessel disease. Human Brain Mapping. https://doi.org/10.1002/hbm.23032

Tullberg, M., Fletcher, E., DeCarli, C., Mungas, D., Reed, B. R., Harvey, D. J., Weiner, M. W., Chui, H. C., & Jagust, W. J. (2004). White matter lesions impair frontal lobe function regardless of their location. Neurology. https://doi.org/10.1212/01.WNL.0000130530.55104.B5

Wang, Z. (2012). Improving cerebral blood flow quantification for arterial spin labeled perfusion MRI by removing residual motion artifacts and global signal fluctuations. Magnetic Resonance Imaging. https://doi.org/10.1016/j.mri.2012.05.004

Wardlaw, J. M., Smith, C., & Dichgans, M. (2019). Small vessel disease: mechanisms and clinical implications. In The Lancet Neurology. https://doi.org/10.1016/S1474-4422(19)30079-1

Wardlaw, J. M., Smith, E. E., Biessels, G. J., Cordonnier, C., Fazekas, F., Frayne, R., Lindley, R. I., O’Brien, J. T., Barkhof, F., Benavente, O. R., Black, S. E., Brayne, C., Breteler, M., Chabriat, H., DeCarli, C., de Leeuw, F. E., Doubal, F., Duering, M., Fox, N. C., … Dichgans, M. (2013). Neuroimaging standards for research into small vessel disease and its contribution to ageing and neurodegeneration. In The Lancet Neurology. https://doi.org/10.1016/S1474-4422(13)70124-8

Watts, D. J., & Strogatz, S. H. (1998). Collective dynamics of ‘small-world9 networks. Nature. https://doi.org/10.1038/30918

Wee, C. Y., Yap, P. T., & Shen, D. (2013). Prediction of Alzheimer’s disease and mild cognitive impairment using cortical morphological patterns. Human Brain Mapping, 34(12), 3411–3425. https://doi.org/10.1002/hbm.22156

Wen, W., Sachdev, P. S., Li, J. J., Chen, X., & Anstey, K. J. (2009). White matter hyperintensities in the forties: Their prevalence and topography in an epidemiological sample aged 44-48. Human Brain Mapping. https://doi.org/10.1002/hbm.20586

Xu, X., Lau, K. K., Wong, Y. K., Mak, H. K. F., & Hui, E. S. (2018). The effect of the total small vessel disease burden on the structural brain network. Scientific Reports, 8(1). https://doi.org/10.1038/s41598-018-25917-4

